# Assessment of Dissemination in Time in Multiple Sclerosis Using a Single 3D T2-FLAIR magnetic resonance imaging Radiomic Signature

**DOI:** 10.64898/2026.10.29.26364762

**Authors:** Elvira G-Guzman, Albert Casajuana-Larqué, Manel Alberich, Luca Bollo, Neus Mongay-Ochoa, Eloy Martínez-Heras, Angela Vidal-Jordana, Xavier Montalban, Mar Tintoré, Àlex Rovira, Jaume Sastre-Garriga, Deborah Pareto

## Abstract

**Background:** In multiple sclerosis (MS), dissemination in time (DIT) is an important component of demonstrating disease activity and establishing the chronology of lesion development. DIT is routinely assessed from contrast-enhanced or follow-up magnetic resonance imaging (MRI). Although proved useful, these methods are not without caveats motivating alternative approaches to study lesion chronology. We investigated whether single T2-FLAIR MRI contains sufficient information to distinguish temporally distinct MS lesion categories using radiomic features.

**Methods:** In a retrospective longitudinal cohort of 273 participants, 39.0 (10.8) y; 172/273 female (63.0%); 101/273 male (37.0%) with MS (1,311 MRI acquisitions), persistent brain lesions were temporally stratified using serial T2-FLAIR MRI. Long-standing established (LS-E) lesions were defined as observed for six years at least, and recent incident (R-I), as lesions that appeared over the span of two years in a follow-up MRI and were present until the last scan available. Radiomic features were extracted from lesion-level T2-FLAIR MRI. Feature selection was performed in an independent discovery cohort (Group 1, N=55). Selected features were then used to train a support vector machine classifier in a separate development cohort with repeated group-wise cross-validation (Group 2, N=163). Final performance was assessed in an independent test cohort (Group 3, N=55). An exploratory analysis was then performed on baseline MRI from 64 subjects with clinically isolated syndrome (CIS), acquired after their first clinical demyelinating event.

**Results:** Group 1 comprised 55 participants with 984 classifiable lesions; paired feature selection was restricted to 32 participants who contributed both lesion categories (658 LS-E and 156 R-I lesions), yielding 18 selected radiomic features. Group 2 comprised 163 participants with 3,203 lesions. In the held-out test cohort, 55 participants contributed 814 lesions (622 LS-E and 192 R-I); the classifier achieved an area under the receiver operating characteristic curve of 0.78 (95% CI, 0.74-0.81). Using the fixed decision threshold, 65% of R-I lesions (125/192; 95% CI, 57%-73%) and 73% of LS-E lesions (457/622; 95% CI, 69%-78%) were correctly classified.

**Conclusions:** Single T2-FLAIR MRI contains measurable information related to MS lesion chronology. This proof-of-concept framework supports the feasibility of a line of research based on radiomics temporal lesion characterisation without contrast administration or dense longitudinal follow-up MRI, with potential relevance for studying lesion evolution and supporting DIT assessment.

## Introduction

Focal brain lesions are central imaging biomarkers in multiple sclerosis (MS), underpinning diagnosis, disease monitoring, and assessment of treatment response (Filippi et al., 2018). Conventional magnetic resonance imaging (MRI), including T1-weighted, T2-weighted, and T2-fluid-attenuated inversion recovery (FLAIR) sequences, enables lesion detection and characterisation in terms of presence, number, and anatomical location, as routinely assessed in clinical MRI reporting. Within the McDonald MS diagnostic criteria framework, lesion patterns have historically been used to demonstrate dissemination of inflammatory demyelination in both space and time, thereby supporting the diagnosis of MS as a chronic disease process rather than a focal monophasic demyelinating event (Thompson et al., 2018). Even if recent revisions to the McDonald criteria reduce the requirement for explicit demonstration of dissemination in time (DIT) in selected diagnostic settings, DIT remains clinically important when the spatial pattern of lesions is less extensive or diagnostically equivocal (Montalban et al., 2025). In such cases, temporal evidence might be required for establishing the diagnosis of MS based on the demonstration of contrast-enhanced lesions or the appearance of new brain lesions on follow-up MRI (Barkhof et al., 2025). This creates a clear opportunity to derive chronological information from routinely acquired MRI, particularly in cases where demonstration of dissemination in space alone is insufficient.

In parallel, growing evidence indicates that MS lesions are not homogeneous entities, but biologically diverse structures with distinct imaging phenotypes, including inflammatory activity, chronic activity, tissue damage, and repair (Lassmann, 2022; Lucchinetti et al., 1996). Imaging markers such as paramagnetic rim lesions, slowly expanding lesions, and different microstructural quantitative MRI measures, suggest that advanced MRI-derived features can capture clinically relevant differences between lesion types (Absinta et al., 2016; Bagnato et al., 2024; Caba et al., 2023; Calvi et al., 2023; Elliott et al., 2019; Smets et al., 2021). Collectively, these approaches reflect a broader shift from lesion detection towards lesion characterisation, in which individual lesions are interpreted not only by their presence and location, but also by the biological information encoded in their imaging phenotype. However, while these developments have advanced the assessment of lesion biology, whether MRI can also capture the chronological state of individual lesions remains less well established (Sweeney et al., 2021).

Currently, lesion chronology is primarily accessible through longitudinal imaging and remains difficult to estimate when MRI sampling is sparse or irregular, as is often the case in routine clinical practice. Moreover, chronological assessment depends on the reliable identification of new lesions by an expert neuroradiologist, a process with wide variability, or on automated tools. In the absence of follow-up MRI, temporal information is largely inferred from gadolinium enhancement, which reflects transient blood–brain barrier disruption during early inflammatory lesion activity and is therefore limited to a short temporal window (Cotton et al., 2003; Saade et al., 2018). Furthermore, repeated administration of gadolinium-based contrast agents is increasingly discouraged, limiting its use as a general marker of lesion chronology (Rovira et al., 2024; Wattjes et al., 2021). Thus, the absence of a widely adopted standardised framework for lesion-level temporal characterisation limits the assessment of lesion chronology in routine clinical practice.

Beyond these practical limitations, chronology is also embedded in any lesion phenotype, yet it is rarely addressed as an independent dimension. For example, the concept of a chronic active lesion combines persistence over time with evidence of ongoing inflammatory activity (Bagnato et al., 2024; Dal-Bianco et al., 2024). Although these dimensions are biologically related, they are not equivalent: lesion chronology refers to the time elapsed since lesion formation, whereas biological state reflects the pathological processes occurring within the lesion. Distinguishing these dimensions provides a framework for potentially isolating the temporal component of lesion evolution from biological activity. Here, we focus specifically on lesion chronology, testing whether conventional MRI contains sufficient information to distinguish long-standing from recently developed lesions irrespective of their biological activity state.

In this study, we investigated whether T2-FLAIR MRI contain sufficient information to infer the relative chronological stage of individual MS lesions. Using longitudinal MRI as a reference, we defined two temporally distinct lesion categories: recently developed lesions and long-standing persistent lesions. We then extracted lesion-level radiomic features and trained a supervised machine-learning classifier to distinguish these chronological categories from a single MRI scan. We evaluated this framework as a proof of concept for single-scan temporal lesion characterisation, with potential relevance for detecting temporally distinct disease activity when gadolinium enhancement or dense longitudinal follow-up is unavailable.

## Materials and Methods

### 1. Cohort and Study Design

This retrospective longitudinal cohort study included participants from a real-world clinical MS cohort at Vall d’Hebron University Hospital, Barcelona, Spain, between 2016 and 2025. Participants underwent a mean (SD) of 5.78 (1.71) MRI examinations over a mean (SD) follow-up of 8.14 (3.17) years; median follow-up was 7.96 years (IQR, 6.58-10.25 years) (see Table 1). All included participants had a final diagnosis of MS according to the 2017 McDonald criteria (Thompson et al., 2018) and at least three MRI acquisitions available on the 3T magnet resulting in a final cohort of 273 eligible participants and a total of 1,311 MRI acquisitions. For the experimental purpose of this work the total cohort was randomly distributed into groups maintaining similar demographic characteristics: Group 1 (G1, n=55, F=34/55 (62%)) with a mean age of 39.2 (10.7) y, Group 2 (G2, n=163, F=103/163 (63.2%)) with a mean age of 39.2 (10.6) y, and Group 3 (G3, n=55, F=35/55 (64%)) with a mean age of 38.4 (11.7) y. Baseline age, recorded sex, and baseline MRI date were complete for all participants in the final analytic cohort. Additionally, we defined the clinically isolated syndrome cohort (CIS) consisting of 64 participants that had their initial 3T baseline MRI acquired within 3-6 months of the CIS event. Participants signed an informed consent. The study was approved by the local Ethical Committee (PR(IDI)625/2021, PR(AG)521/2022). No a priori sample-size calculation was performed. Study size was determined by the number of participants in the retrospective cohort who met the prespecified eligibility and imaging-availability criteria.

**Table 1.** Participant, longitudinal MRI, and lesion characteristics by analysis group.

| **Characteristic** | **Overall (N=273)** | **Group 1 Discovery (n=55)** | **Group 2 Development (n=163)** | **Group 3 Held-out Test (n=55)** |
|---|---|---|---|---|
| Participants, No. | 273 | 55 | 163 | 55 |
| Age, mean (SD), y | 39.0 (10.8) | 39.2 (10.7) | 39.2 (10.6) | 38.4 (11.7) |
| Sex, No. (%) |  |  |  |  |
| Female | 172 (63.0%) | 34 (62%) | 103 (63.2%) | 35 (64%) |
| Male | 101 (37.0%) | 21 (38%) | 60 (36.8%) | 20 (36%) |
| MRI examinations per participant, mean (SD) | 5.78 (1.71) | 5.58 (1.64) | 5.87 (1.70) | 5.71 (1.81) |
| Follow-up duration, mean (SD), y | 8.14 (3.17) | 7.92 (3.42) | 8.16 (2.97) | 8.32 (3.50) |
| Follow-up duration, median (IQR), y | 7.96 (6.58-10.25) | 8.16 (6.44-10.02) | 7.83 (6.83-10.23) | 8.28 (6.24-10.74) |
| Consecutive interscan interval, mean (SD), y | 1.70 (1.32) | 1.73 (1.34) | 1.68 (1.30) | 1.77 (1.34) |
| Consecutive interscan interval, median (IQR), y | 1.12 (0.97-2.02) | 1.10 (0.95-2.12) | 1.13 (0.97-1.97) | 1.11 (0.99-2.07) |
| Classifiable lesions, No. | 5001 | 984 | 3203 | 814 |
| LS-E lesions, No. (%) | 3778 (75.5%) | 729 (74.1%) | 2427 (75.8%) | 622 (76.4%) |
| R-I lesions, No. (%) | 1223 (24.5%) | 255 (25.9%) | 776 (24.2%) | 192 (23.6%) |

**Abbreviations:** IQR, interquartile range; LS-E, long-standing established; MRI, magnetic resonance imaging; R-I, recent incident; SD, standard deviation.
**Notes:** Age and sex were complete for all 273 participants. Interscan summaries are based on consecutive MRI intervals (overall n=1305). MRI examinations per participant reflect longitudinal time points available for lesion tracking; radiomics analyses used 3D FLAIR only.

### 2. MRI Acquisition Protocol and Preprocessing

The MRIs were acquired in a 3T Prisma magnet (Siemens, Erlangen, Germany). The protocol included a sagittal 3D T1 magnetization prepared rapid gradient-echo (MPRAGE) (TR = 2300 ms, TE = 2.98 ms; flip angle = 9°; 176 slices, voxel size = 1.0 x 1.0 x 1.0 mm3) and 2D T2-weighted fluid attenuation inversion recovery (FLAIR) (TR = 9000 ms, TE = 93 ms, TI = 2500 ms, flip angle = 120°, voxel size = 0.49 x 0.49 x 3.0 mm3) or 3D T2-weighted FLAIR (TR = 7000 ms, TE = 410 ms, TI = 2050 ms, voxel size = 1.0 x 1.0 x 1.0 mm3). Across all MRI acquisitions, T2-FLAIR imaging was predominantly performed in 3D (83.1%). Radiomic feature extraction and all subsequent radiomics-based analyses were performed exclusively on 3D FLAIR images, 2D FLAIR lesion masks were used only for longitudinal lesion tracking.

All MRIs were reoriented to a standard anatomical orientation. Intensity inhomogeneities were corrected using N4 bias field correction (Tustison et al., 2010). Then, a robust field-of-view normalisation was applied to reduce variability related to differences in T1-weighted brain coverage across acquisitions. For each participant and acquisition, the T2-FLAIR was co-registered to the T1-weighted MRI. All follow-up MRIs were registered (affine transformation) to baseline MRI, which served as the within-participant reference space. The same transformation was then applied to the corresponding T2-FLAIR MRI, ensuring consistent alignment of both modalities at baseline space (Figure 1, panel A). These steps were performed using the FMRIB Software Library (FSL) (Jenkinson et al., 2012).

**Figure 1.**
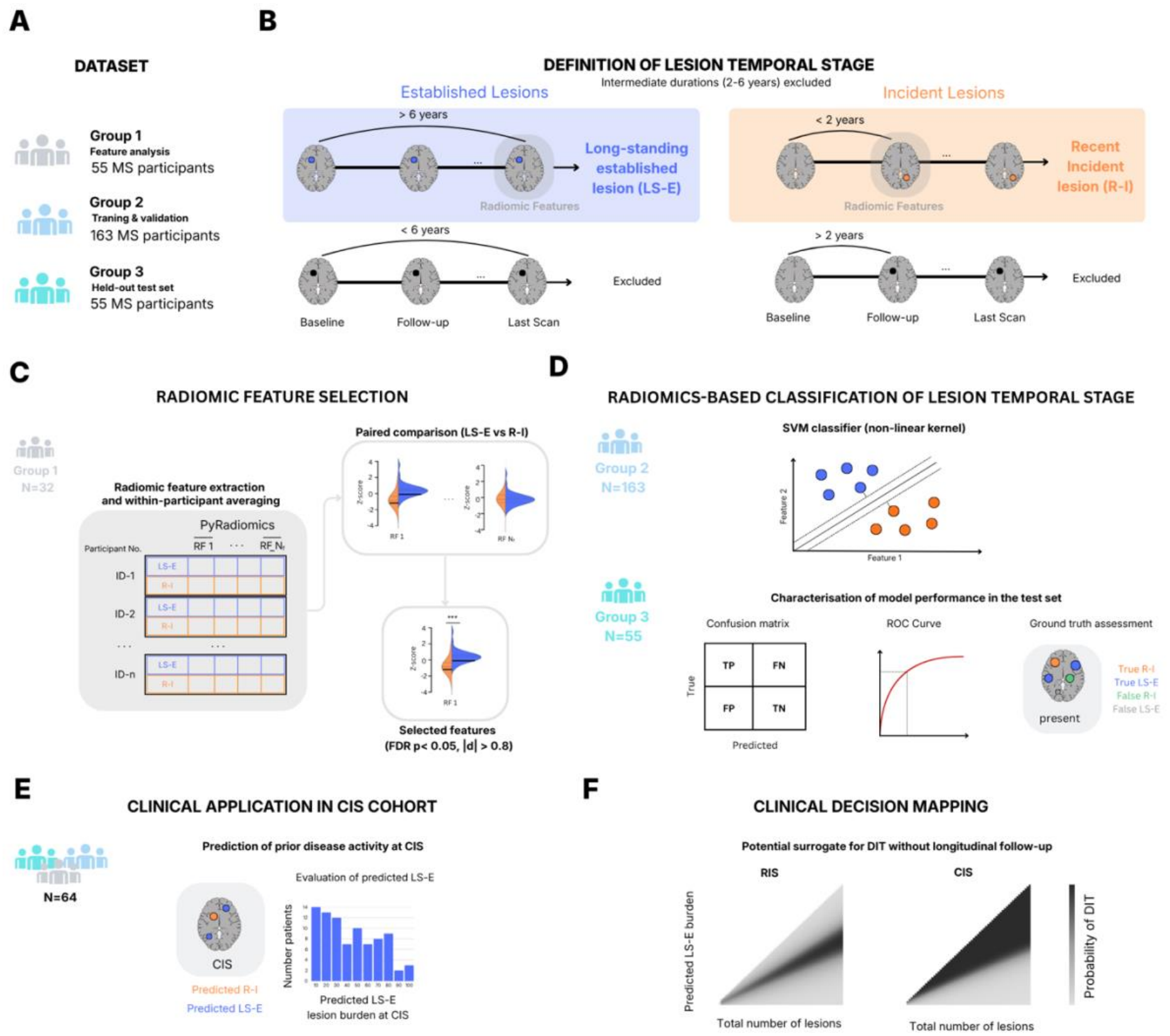
Radiomics-based classification of lesion temporal stage and clinical application. **(A)** Study design and dataset composition, including feature analysis (Group 1), training/validation (Group 2), and held-out test set (Group 3). **(B)** Definition of lesion temporal stages based on longitudinal MRI: long-standing established lesions (LS-E, >6 years) and recent incident lesions (R-I, <2 years). Lesions not meeting the criteria were excluded. **(C)** Radiomic feature extraction and selection pipeline. Features were averaged within participants, followed by paired comparison between LS-E and R-I lesions. Significant features were selected based on FDR-adjusted (P < .05) and effect size (|d| > 0.8). **(D)** Radiomics-based classification using a support vector machine (SVM). Model performance was evaluated in an independent test set using confusion matrix and ROC analysis. **(E)** Clinical application in a CIS cohort (n = 64), showing estimation of predicted LS-E lesion burden. **(F)** Clinical decision mapping framework integrating lesion count and predicted LS-E burden to estimate the probability of dissemination in time (DIT), with potential application in RIS and CIS settings.

### 3. Lesion Tracking and Temporal Category Definitions (LS-E vs R-I)

White matter lesions were segmented at individual acquisition level using the Lesion Segmentation Toolbox-Artificial Intelligence (LST-AI) (Wiltgen et al., 2024). The lesion map was thresholded (0.3), binarised, and individual lesions were separated using connected component analysis. For each lesion, geometric properties including volume and centroid location were computed. Lesions smaller than 15 voxels (15 mm3) were excluded from the analysis (Elliott et al., 2019).

MRI follow-up reflected routine clinical practice, resulting in heterogeneous but broadly comparable longitudinal sampling across groups (Suppl. mat. Figure 1, panel B). Lesions were matched across acquisitions in a common baseline space using predefined spatial proximity and overlap criteria (maximum centroid distance 5 mm and/or >10% lesion overlap), with all correspondences visually confirmed. For each lesion, first detection was defined as the earliest MRI acquisition in which the lesion was identified. Lesion status was also assessed at the participant’s last available MRI, and lesions not detectable at final follow-up were excluded from further analysis.

Lesions were initially classified according to their first appearance during longitudinal MRI follow-up. Lesions already present at first MRI available and persisting to the final available MRI were labelled as Established, whereas lesions that appear on a follow-up MRI and persisting to the final available scan were labelled as Incident. Established lesions were further classified as Long-standing Established (LS-E) when the interval between baseline and final MRI exceeded six years. Incident lesions were classified as Recent Incident (R-I) when the interval between lesion detection and the preceding MRI was less than two years. Lesions not meeting these temporal criteria were excluded to minimise uncertainty in lesion chronology. Missing radiomic feature values were imputed using median values estimated from the training data and applied unchanged to the corresponding validation or test data.

### 4. Radiomic Feature Selection (Group 1)

A total of 103 lesion-level radiomic features, together with distance to the centre of the brain, were extracted from T2-FLAIR images using PyRadiomics (van Griethuysen et al., 2017). Features were computed from individual lesion masks and included first-order intensity, shape, and texture descriptors derived from from gray-level co-occurrence, gray-level run-length, gray-level size-zone, and gray-level dependence matrices. Intensity normalisation was applied prior to feature extraction, and texture features were computed using a fixed bin width of 5. Lesions smaller than 200 voxels were isotropically dilated before texture computation to improve numerical stability in small lesions (Zwanenburg et al., 2020).

Feature selection was performed exclusively in Group 1, which served as the independent discovery cohort. To enable within-participant comparisons, only participants presenting both LS-E and R-I lesions were included. Radiomic features for LS-E lesions were extracted from the participant’s last available MRI, whereas features for R-I lesions were extracted from the MRI in which the lesion was first detected. Group 1 included 55 participants and 984 classifiable lesions. The paired feature-selection analysis was restricted to 32 participants who contributed both LS-E and R-I lesions, comprising 658 LS-E and 156 R-I lesions before participant-level averaging. Thus, the paired statistical comparisons were based on 32 participant-level pairs. Multiple comparisons were controlled using false discovery rate (FDR) correction, with FDR-adjusted P < .05 considered significant. Features were retained when they satisfied both statistical significance and a large effect size threshold (absolute Cohen’s d > 0.8). Selected features were subsequently used for machine learning classification of lesion temporal category (Cortes & Vapnik, 1995).

### 5. Model Development (Group 2) and Held-out Test Evaluation (Group 3)

Radiomic features selected in the discovery stage were used to train a supervised classifier for distinguishing LS-E from R-I lesions. Model development was performed exclusively in Group 2. A support vector machine classifier with radial basis function kernel (SVM) was trained at the lesion level, with each lesion treated as an independent input to the model. To prevent data leakage, data partitioning was performed at the participant level, ensuring that all lesions from a given participant were assigned exclusively to either training or validation sets. Class imbalance was addressed using cost-sensitive learning, with class weights inversely proportional to class frequencies during SVM optimisation. To assess feature collinearity and the potential influence of unequal lesion burden across participants, complementary sensitivity analyses were performed.

Model performance in Group 2 was estimated using repeated five-fold group cross-validation (5 folds × 10 repetitions), with participant-level partitioning to ensure that lesions from the same participant were never split across training and validation folds. Within each training fold, we used feature standardisation, with parameters derived exclusively from the training data and applied unchanged to the validation set. Model performance was assessed using receiver operating characteristic (ROC) analysis and summarised by the area under the curve (AUC). Cross-validated predictions were pooled to derive confusion matrices and standard performance metrics, including precision, recall, F1-score, and accuracy. The operating decision threshold was determined in Group 2 by maximising the Youden index on pooled cross-validated predictions (Youden, 1950).

Held-out performance was quantified using the area under the receiver operating characteristic curve (AUC) and threshold-dependent classification metrics. The decision threshold was determined exclusively in Group 2 by maximizing the Youden index on pooled cross-validated predictions and was applied unchanged to Group 3. Because multiple lesions were contributed by the same participant, 95% CIs for held-out performance metrics were estimated using a participant-cluster bootstrap with 5,000 resamples, in which participants were sampled with replacement and all lesions from each sampled participant were retained. As a sensitivity analysis, test lesions were weighted so that each participant contributed equal total weight, with each lesion receiving weight equal to the inverse of the number of test lesions contributed by that participant. A participant-cluster bootstrap was also used to estimate the 95% CI for the weighted AUC.

Model interpretability was assessed using SHAP values (Lundberg & Lee, 2017), computed on a stratified subset of Group 2 to reduce computational burden while preserving class balance. SHAP analysis quantified the contribution of individual radiomic features to lesion-level predictions and was summarised using SHAP summary plots, where positive values indicate contributions towards LS-E classification and negative values towards R-I classification.

Following cross-validation, a final SVM model was trained using all available Group 2 lesion-level data and subsequently evaluated in the independent test cohort (Group 3). To facilitate participant-level interpretation, lesion-level predictions obtained using the fixed decision threshold were mapped back onto MRI lesion masks and compared with longitudinally derived ground-truth lesion categories. To reduce temporal-label uncertainty, lesions with intermediate or insufficiently sampled temporal histories were excluded. Discovery, model development, and held-out testing were performed in nonoverlapping participant groups, and participant-level grouping was maintained during cross-validation. Class weighting addressed lesion-class imbalance, and an inverse-lesion-contribution sensitivity analysis assessed the influence of participants with high lesion burden.”

### 6. Exploratory Analyses

#### 6.1 Patient-Level Clinical application at CIS & Bayesian DIT framework

An exploratory analysis was performed to assess lesion chronology at the CIS cohort baseline MRI. Indeed, the first available MRI scan coincides with the first clinical event. Because the classifier had been trained exclusively on follow-up and final MRIs, baseline scans represented entirely unseen data and could therefore be analysed across participants from all study groups without data leakage. Only participants whose baseline MRI corresponded to the first clinical event were included, resulting in a final cohort of 64 participants. The acquisition protocol was identical to that describe in Section 2, nonetheless baseline MRIs were acquired on a 3.0 T Trio Siemens magnet, which was subsequently upgraded to a Prisma.

An exploratory probabilistic framework was developed to estimate DIT at the patient level using lesion-level classifier outputs. For each participant, the framework considered the total number of lesions (N) and the number classified as LS-E-like (k) using the fixed lesion-level decision threshold. Posterior probabilities of DIT were modelled as a function of these variables, combining the likelihood of observing k LS-E-like lesions given N with scenario-dependent prior probabilities in patients presenting with radiologically isolated syndrome (RIS) and CIS:

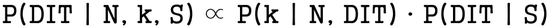

The likelihood term was defined to increase monotonically with the number of LS-E-like lesions, reflecting the assumption that a higher burden of LS-E-like lesions provides stronger evidence of prior disease activity. Posterior probabilities were computed across a range of lesion counts and visualised as two-dimensional heatmaps for Radiological Isolated Syndrome (RIS) and CIS scenarios, as they are both clinical situations that would deeply benefit from an objective estimation of lesion chronology. This framework provides an interpretable mapping from lesion-level temporal classifications to patient-level DIT estimates, without implying a calibrated clinical prediction model. In the CIS setting, DIT was operationalised as the presence of at least one true LS-E-like lesion. For an individual exploratory analysis of DIT at CIS cohort posterior probabilities were computed at the participant level and used to stratify participants into four confidence categories: DIT unlikely, defined as posterior probability <0.50; possible DIT, 0.50–0.80; probable DIT, 0.80–0.95; and highly probable DIT, ≥0.95. Meanwhile as a RIS scenario suppose a person that presents lesions on MRI but no symptomatology, to support a MS diagnosis we need to proof the existence of both LS-E-like lesions and R-I-like lesions in order to demonstrate lesion activity in different moments of time, estiblish a positive DIT.

#### 6.2 Exploratory Longitudinal Analysis of Lesion Evolution

An exploratory longitudinal analysis was performed to assess whether the classifier captured imaging features related to lesion evolution over time. Specifically, we evaluated whether known R-I lesions showed increasing probabilities of LS-E classification with increasing lesion age. Only lesions with at least two follow-up MRIs after first detection were included. We evaluated lesions that were not part of the training or validation of the model. For each lesion, radiomic features were extracted at each available follow-up MRI and processed using the trained classifier. Lesion age was defined as the time elapsed since first detection. Predicted probabilities were aggregated using adaptive time bins, and mean LS-E probabilities were computed to visualise longitudinal trends. Because lesion-level chronological ground truth was not available at baseline, LS-E-like and R-I-like predictions should be interpreted as radiomic profiles compatible with different temporal lesion states rather than as exact lesion-age estimates.

## Results

### Study design and temporally stratified lesion cohort

The analytical workflow is summarised in Figure 1. Across the 273 participants 39.0 (10.8) y; 172/273 female (63.0%); 101/273 male (37.0%), the full discovery group (Group 1) contained 984 classifiable lesions, the model-development group (Group 2) contained 3,203, and the held-out test group (Group 3) contained 814. LS-E lesions accounted for 729 of 984 lesions (74.1%) in Group 1, 2,427 of 3,203 (75.8%) in Group 2, and 622 of 814 (76.4%) in Group 3. Additional details on cohort composition, longitudinal sampling, and lesion categorisation are provided in Supplementary Figures 1 and 3.

### Radiomic differences and lesion temporal classification

Feature selection was performed in 32 Group 1 participants who contributed both lesion categories. Eighteen radiomic features met both predefined criteria, with absolute paired Cohen d values ranging from 0.83 to 1.65 and all FDR-adjusted P < .001 (Figure 2A; Supplementary Table 3). These included first-order intensity features (Maximum, Mean, Root Mean Squared, 90 Percentile, 10 Percentile, and Median), shape descriptors (Sphericity, Major Axis Length, Minor Axis Length, Elongation, Flatness, and maximum lesion diameters), and run-length texture features (Run Variance, Run Length Non-Uniformity Normalized, and Run Entropy). The selected features were then used to train a SVM classifier in Group 2, comprising 163 participants with 2,427 LS-E and 776 R-I lesions. Group 3 comprised 55 participants contributing 814 lesions (622 LS-E and 192 R-I). In this held-out test cohort, the classifier achieved an AUC of 0.78 (95% CI, 0.74-0.81). Using the fixed decision threshold selected in Group 2, 125 of 192 R-I lesions (65.1%; 95% CI, 57.0%-73.5%) and 457 of 622 LS-E lesions (73.5%; 95% CI, 68.5%-77.9%) were correctly classified (Figure 2, panel B-D). Cross-validation performance during model development is shown in Supplementary Figure 2. Discrimination was similar when participants were weighted inversely by their lesion contribution (participant-weighted AUC, 0.77; 95% CI, 0.72-0.82; Supplementary Table 2).

**Figure 2.**
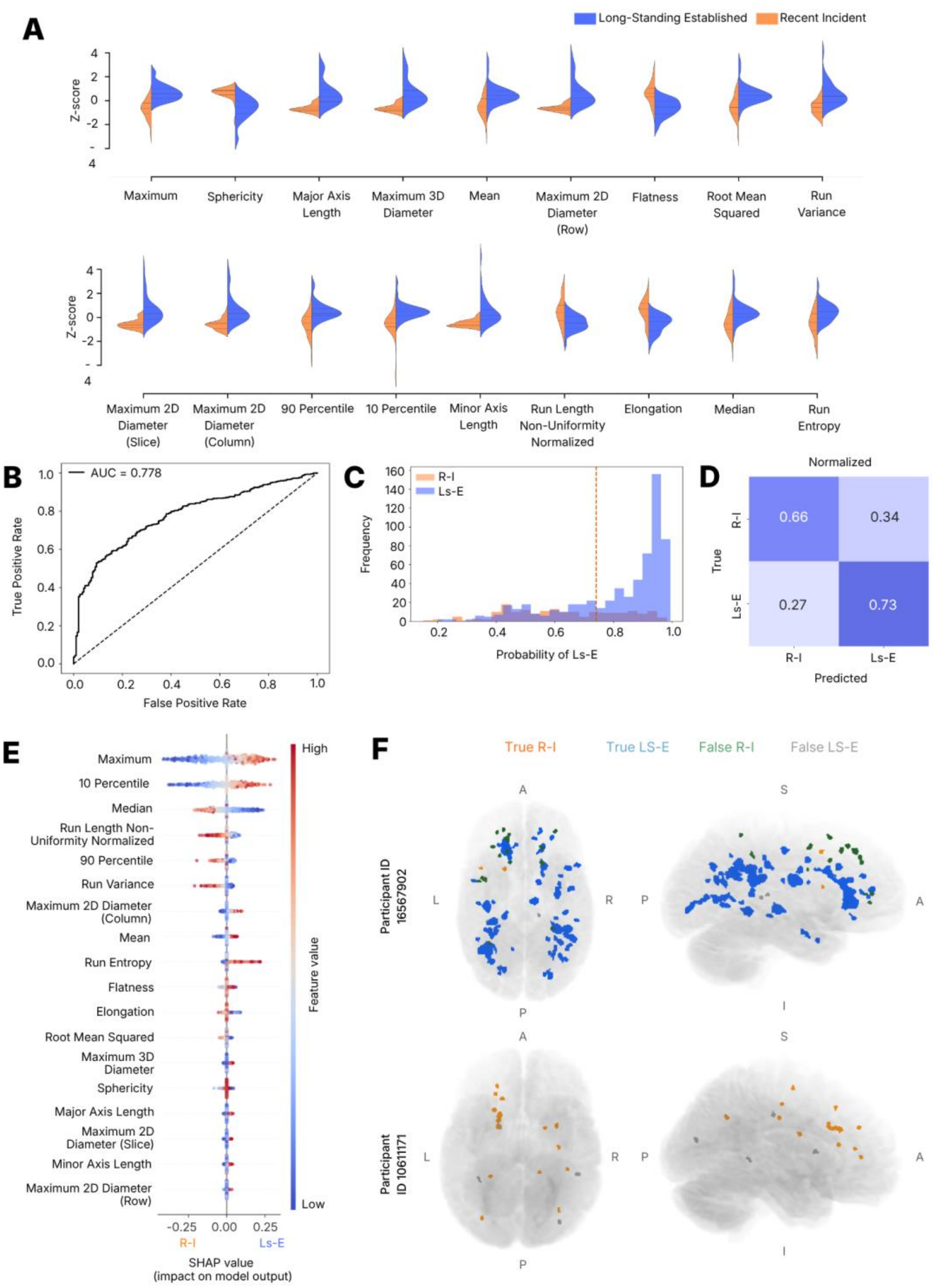
Radiomic feature differences, classification performance, and model interpretability. **(A)** Distribution of selected radiomic features (Z-scores) comparing long-standing established lesions (LS-E, blue) and recent incident lesions (R-I, orange). Features include shape and texture descriptors derived from lesion masks. **(B)** Receiver operating characteristic curve for the held-out test cohort (AUC, 0.78; 95% CI, 0.74-0.81). The 95% CI was estimated using participant-cluster bootstrap resampling. **(C)** Distribution of predicted probabilities for LS-E across lesions, showing separation between R-I (orange) and LS-E (blue). The dashed vertical line indicates the classification threshold. **(D)** Normalised confusion matrix summarizing classification performance, with proportions of correctly and incorrectly classified lesions for each class. **(E)** SHAP (SHapley Additive exPlanations) summary plot illustrating feature importance and directionality of effects on model output. Each point represents a lesion, coloured by feature value (low to high), with horizontal position indicating impact on classification toward LS-E or R-I. **(F)** Representative spatial maps of lesion-level predictions in two participants. Lesions are colored according to classification outcome: true R-I (orange), true LS-E (blue), false R-I (green), and false LS-E (grey), demonstrating spatial distribution and model performance at the individual level.

To further interpret model behaviour, SHAP analysis was performed on the final classifier (Figure 2, panel E). The highest-ranking contributors were predominantly intensity- and texture-related features, with Maximum, 10 Percentile, Median, Run Length Non-Uniformity Normalized, 90 Percentile, Run Variance, and Run Entropy showing the greatest overall impact on model output. Shape descriptors were also represented among the informative features, but their contribution appeared less prominent overall. Representative lesion maps from the independent test cohort further illustrate the spatial distribution of correct and incorrect lesion-level classifications within individual participants (Figure 2, panel F).

### Clinical application at CIS

Predicted lesion temporal composition at CIS showed marked inter-patient heterogeneity (Figure 3, panel A-B). Most participants had several lesions with a LS-E-like profile at baseline, although the proportion and absolute burden of such lesions varied widely across individuals. The distribution of LS-E-like lesion burden by both volume and count is shown in Supplementary Figure 4. Finally, illustrative proof-of-concept patient-level probability maps showed that the estimated likelihood of DIT increased with both the number and proportion of LS-E-like lesions in both CIS and RIS scenarios (Figure 3, panel D-E).

**Figure 3.**
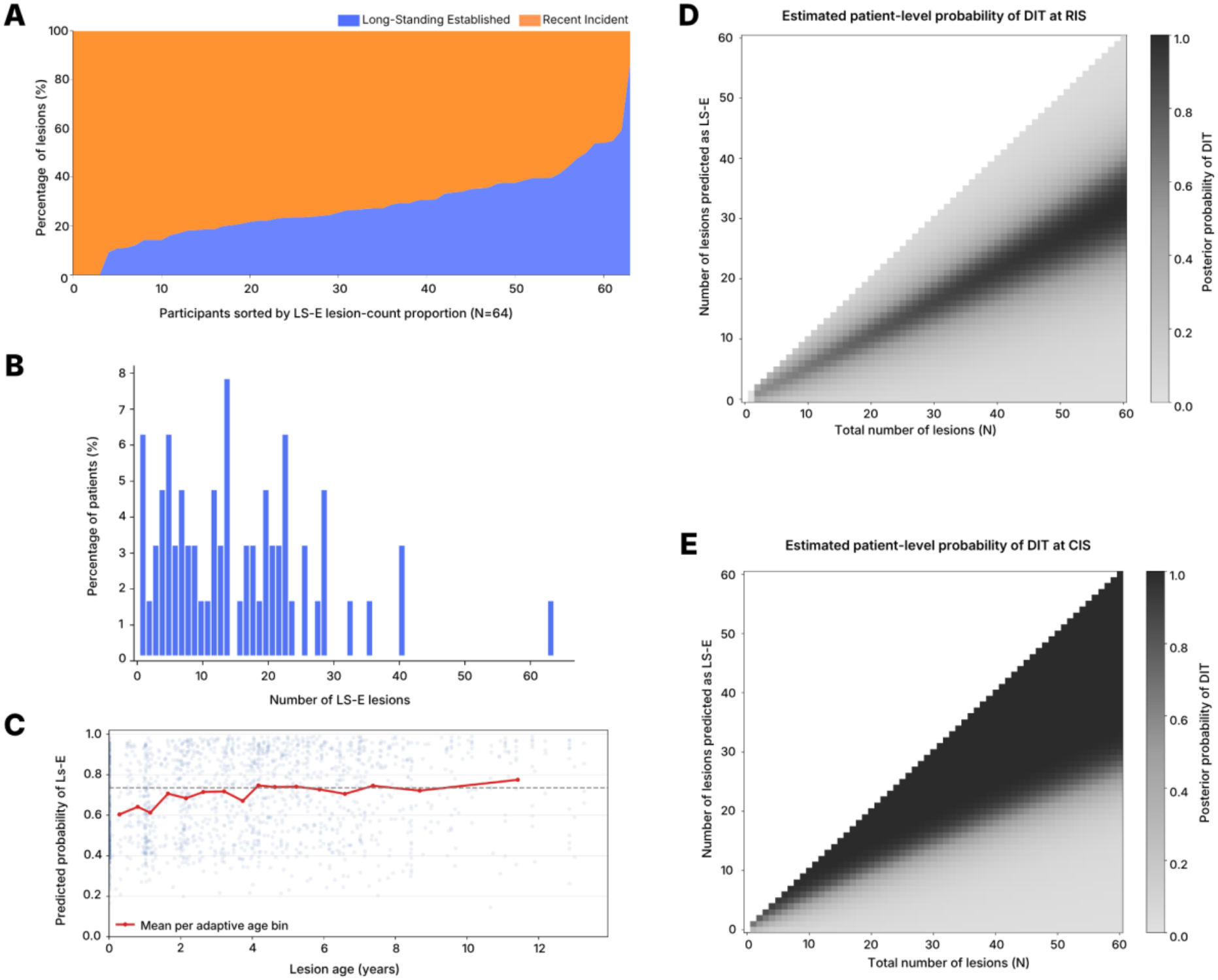
Clinical relevance of radiomics-derived lesion temporal staging and patient-level decision mapping. **(A)** Distribution of the proportion of lesions classified as long-standing established (LS-E, blue) and recent incident (R-I, orange) at baseline in the CIS cohort (N = 64), with participants ordered by increasing LS-E proportion. **(B)** Histogram showing the distribution of the absolute number of LS-E lesions per patient at baseline. **(C)** Relationship between predicted probability of LS-E and lesion age. Each point represents a lesion, with the red line indicating the mean probability across adaptive age bins, demonstrating increasing probability of LS-E classification with lesion age. **(D)** Estimated patient-level posterior probability of dissemination in time (DIT) in a radiologically isolated syndrome (RIS) scenario, as a function of total lesion count and number of lesions classified as LS-E. **(E)** Equivalent decision mapping for the CIS scenario.

To translate lesion-level temporal classifications into a patient-level estimate we used an equal-odds reference prior, assigning the hypotheses of DIT and no DIT an initial probability of 0.50 each. This reference prior should not be interpreted as an estimate that 50% of CIS patients truly satisfy DIT; rather, it provides a transparent and neutral starting point from which the observed lesion composition can modify the probability of containing at least one true LS-E lesion. Under this framework, 6 of 64 participants (9.4%) reached a posterior probability of at least 0.80, and 5 of 64 (7.8%) reached a posterior probability of at least 0.95. Thus, in approximately one in ten participants, the baseline lesion composition was sufficiently unusual that it was difficult to explain solely through the expected false-positive behaviour of the lesion classifier.

### Exploratory longitudinal analysis of lesion evolution

In support of the idea that age could shift the lesion phenotype from R-I-like to LS-E-like we ran an exploratory longitudinal analysis of known R-I lesions. The probability of LS-E classification increased with lesion age, supporting the interpretation that the classifier captures features related to lesion evolution over time rather than a fixed binary separation alone (Figure 3, panel C). Lesions of four years onwards, on average show a probability of 80% for LS-E phenotype, that is the probability threshold used in the present study.

## Discussion

This study provides, to our knowledge, the first proof-of-concept evidence that single 3D T2-FLAIR MRI can distinguish between temporally distinct classes of MS lesions without the need of contrast administration or serial MRI. Using a large, well-characterised longitudinal MRI cohort, we define widely separated temporal reference categories and found that R-I and LS-E lesions showed distinct radiomic profiles, and the classifier achieved an AUC of 0.78 (95% CI, 0.74-0.81) in the held-out test cohort. Classification was asymmetric across lesion categories, with correct classification of 65.1% of R-I lesions and 73.5% of LS-E lesions. In fact, existing lesions at the first clinical presentation present radiomic features compatible with both long term and recent categories, highlighting that lesion composition is heterogeneous and subject specific. These findings suggest that lesion chronology leaves a measurable imprint on T2-FLAIR MRI, and that this knowledge could be useful to demonstrate DIT aiding an earlier diagnosis.

Radiomics has increasingly been used to characterise MS lesions beyond their visual appearance, often to classify lesion activity, subtype, clinical phenotype, or subsequent evolution. The literature describe discriminative information being distributed across combinations of features such as intensity, shape, and texture descriptors (Alomair et al., 2025; Elhaie et al., 2025; Lambe et al., 2025; Rostami et al., 2024; Shi et al., 2024). Our findings are consistent with this multidimensional view of lesion characterisation, the temporal separation between LS-E and R-I lesions on T2-FLAIR MRI is expressed through concurrent changes in signal distribution, lesion morphology, and internal textural organisation. Interestingly, there is a substantial overlap between the specific features identified in our study and the ones previously described in the characterisation of different MS lesion types. Rostami et al. identified Maximum, Median, Elongation and Sphericity among the most informative features for distinguishing active from non-active lesions (Rostami et al. 2024), all of which were also selected in our temporal comparison. Similarly, Shi et al. reported first-order measures including the 10th percentile, Median, Mean and Root Mean Squared intensity, together with texture descriptors of grey-level heterogeneity, as discriminative across different lesion phenotypes (Shi et al., 2024). Other studies have likewise emphasised the contribution of FLAIR-derived texture features to lesion activity classification (Elhaie et al., 2025). This convergence suggests that lesion chronology is reflected in the same broad dimensions of tissue signal, morphology, and internal heterogeneity that distinguish other biologically meaningful lesion states. Our findings extend previous results by showing that these radiomic dimensions also contain information about relative lesion chronology from a single non-contrast T2-FLAIR MRI.

Beyond T2-FLAIR, MRI sequence of choice also varies across the literature; while some studies derive radiomic signatures from a single conventional sequence (Alomair et al., 2025; Rostami et al., 2024; Shi et al., 2024), others combine information from several macro and microstructural MRI biomarkers, with different sequences contributing distinct radiomic profiles (Elhaie et al., 2025; Sweeney et al., 2021). Even in the presence of multiple sequences, Elhaie et al. reported an important contribution of T2-FLAIR-derived features within a multisequence model. Indeed, they found other features such as lesion location had limited influence on classification performance (Elhaie et al., 2025), which is consistent with our results. Similarly, Peng et al. showed that T2-FLAIR radiomics could predict subsequent enlargement or shrinking of unenhanced MS lesions, further supporting the view that T2-FLAIR contains quantitative information related to lesion evolution (Peng et al., 2021).

Having established that temporally distinct lesion categories differed across several radiomic domains, analysing our classifier clarify how these differences contributed to prediction. SVM operates in a multidimensional feature space, in which classification reflects the combined contribution of partially overlapping radiomic descriptors. SHAP analysis suggested that this joint signature was driven primarily by intensity- and texture-related features, whereas geometric descriptors contributed less prominently overall. These features should not be interpreted as fully independent determinants of classification, given the partial collinearity between radiomic variables, we should think of them as representative markers of broader differences in lesion intensity distribution and intralesional texture.

Although much work has been done in lesion characterisation, lesion age itself has rarely been the explicit target of MS radiomics studies. Sweeney et al. directly investigated this question using a multisequence MRI-based from single-timepoint imaging (Sweeney et al., 2021). Their approach estimated continuously lesion age within a 0-to 4-year window using first-order radiomic features derived from multiple MRI sequences, including T1-weighted, T2-weighted, T2-FLAIR, contrast-enhanced T1-weighted imaging, and quantitative susceptibility mapping, together with demographic information (Sweeney et al., 2021). Our findings therefore extend this effort by studying temporal lesion classification rather than a continuous estimation of their age using only clinically acquired T2-FLAIR MRIs, in a larger cohort and moving beyond first order radiomic features. Their best-performing model integrated information across all sequences, with contrast-enhanced T1-weighted features playing a crucial role in identifying early lesions, given that gadolinium enhancement act as a direct marker of the acute inflammatory phase. Importantly, however, T2-weighted features contributed among the most important predictors when modelling lesion age beyond the initial acute stage. In fact, T2-weighted mean intensity was the most important feature in their simplified two-step model based on mean signal intensity alone. Consistent with our findings, temporal information is retained within T2-sensitive lesion signal rather than being exclusively dependent on acute contrast enhancement or specialised quantitative sequences. Our study extends this previous work by demonstrating temporal lesion stratification using T2-FLAIR alone, across substantially larger lesion and participant samples and over more widely separated temporal categories, while incorporating first-order, morphological and textural radiomic information rather than first-order features alone.

Even though our results show a good classification accuracy, it is worth to address the classification asymmetry between lesion categories. LS-E lesions were identified somewhat more accurately than R-I lesions despite explicit handling of class imbalance during training. One possible explanation is that LS-E lesions represent a more stable and internally consistent imaging phenotype, whereas lesions within the first two years after appearance may encompass greater radiological heterogeneity and more dynamic tissue change, thereby reducing separability within the recent-lesion category. This asymmetry suggests that the radiomic signature of lesion chronology may become more stable with time, whereas recently formed persistent lesions may occupy a more variable transitional state.

The exploratory analysis on the CIS cohort provides a first indication of how lesion-level temporal classification might be translated into patient-level clinical information. More than 50% of CIS cohort participants already showed at least some lesions with a LS-E-like profile at first clinical presentation. Notwithstanding, the absolute and proportional burden of such lesions varied widely, suggesting that the first clinical event may occur against different backgrounds of prior, clinically silent lesion accumulation at temporally different scenarios rather than simply different amounts of total lesion load. This interpretation is consistent with a broader perspective in which inflammatory demyelinating disease may begin before the first recognised clinical event. In fact MS prodrome, presymptomatic MS, and RIS, may serve as examples of this view emphasising that asymptomatic MRI abnormalities can represent biologically meaningful disease activity and can contribute to estimating the risk of clinical conversion (Giovannoni, 2017; Lebrun-Frénay et al., 2023; Montalban et al., 2025; Okuda & Lebrun-Frénay, 2024).

From this perspective, radiomics-derived temporal profiling from a single baseline T2-FLAIR MRI could eventually provide a supportive marker of prior lesion accumulation and help address one of the central diagnostic pillars in MS, demonstrating DIT. This is clinically important because evidence of DIT often relies on contrast enhancement or the appearance of new lesions on follow-up T2-weighted MRI, approaches that are useful but have known limitations in early diagnostic pathways. Although according to clinical guidelines gadolinium use at the onset of the disease is recommended, the age estimation of the lesions is restricted to 3-4 weeks, and many non-enhancing lesions are observed, and waiting for new lesions to appear may delay diagnosis and treatment in a disease where earlier intervention is intended to reduce future inflammatory activity and irreversible tissue damage (Ramalho et al., 2016). Patients presenting with CIS, radiomic temporal profiling could complement their diagnostic by helping to identify whether the baseline lesion load already contains imaging evidence compatible with previous lesion events. The same information would have a related but distinct role in RIS, where radiomic evidence of lesions occupying different temporal states could support the inference that demyelinating activity has occurred at more than one time point. Such information would be relevant and could impact surveillance intensity, MRI timing, or treatment management. Importantly, revised RIS criteria and recent randomised trials suggest that earlier identification and intervention in high-risk individuals decreases the time to a first clinical event (Lebrun-Frénay et al., 2023; Lebrun-Frenay et al., 2025; Okuda et al., 2023; Okuda & Lebrun-Frénay, 2024). The patient-level DIT probability maps should therefore be viewed not merely as illustrative outputs, but as proof-of-concept examples of how the same lesion-level temporal information might be translated differently according to clinical context, supporting diagnostic reasoning in CIS and risk-oriented monitoring in RIS. Further validation, calibration, and prospective testing will be required before such estimates can be considered diagnostic or prognostic tools.

Once temporal lesion classification is possible, even within broad temporal categories, it opens questions about the hidden chronology of the lesion burden, not only about diagnosis. The presence of LS-E-like lesions at CIS raises the possibility that the first clinical event may occur against a variable background of prior clinically silent lesion load and invites future work to examine whether lesions associated with symptomatic presentation follow the same temporal and phenotypic trajectory as lesions that remain clinically silent. More precise lesion-age estimation, building on the approach introduced by Sweeney et al., could help clarify how lesion formation, lesion persistence, and clinical onset relate to one another (Sweeney et al., 2021). In that sense, chronology may become not only a diagnostic criterion to be satisfied, but rather an identifiable dimension of lesion characteristic. Biological features intersect with the age of a lesion, and the latter may be overlooked if we consider lesions only through biologically defined categories (i.e. remyelinated, chronic-active, slowly evolving…). A related point is that the present analysis deliberately focused on lesions that remained visible over time, in other words, they were persistent. This was necessary to define temporally anchored lesion categories and to compare radiomic profiles within lesions that could be reliably followed across longitudinal MRI. They should not, however, be taken to represent the full biology of lesion evolution, which also includes lesions that resolve, fluctuate, or continue to change slowly over time (Ciccarelli et al., 1999; Elliott et al., 2019; Meier et al., 2007; A. Rovira et al., 2013; Sethi et al., 2017). Whether these different trajectories carry distinct temporal and radiomic signatures remains an important question for future work.

This study is not without limitations, because lesion onset could not be determined precisely from retrospective clinical follow-up with heterogeneous scan intervals, continuous lesion-age modelling was not feasible within the present dataset. We therefore defined deliberately separated temporal categories to reduce temporal ambiguity, while recognising that this constrained the scope of inference. Similarly, the focus on lesions that remained detectable over time reflected the need for a stable longitudinal imaging substrate, meaning that the results apply primarily to the persistent component of the lesion burden. Future validation across sites, scanners, acquisition protocols, and segmentation pipelines will therefore be needed to determine how robustly these findings generalise beyond the present setting. The present framework also relied on an intentionally interpretable machine learning approach based on predefined radiomic features and SVM. More advanced models trained on larger and more densely sampled longitudinal MRI datasets may eventually allow finer temporal stratification and more precise lesion-age estimation, particularly if temporal imaging information is integrated with biological lesion markers and clinical outcomes. The exploratory CIS, RIS, and DIT analyses should therefore be viewed as hypothesis-generating rather than clinically validated. Nevertheless, the present findings provide an encouraging proof of concept that temporally distinct persistent MS lesions can be differentiated using a single T2-FLAIR MRI. The present findings suggest that conventional MRI holds measurable chronological information about MS lesions, opening a path towards deeper understanding of lesion biology and, with further development, earlier diagnosis in MS.

## Data Availability

All data produced in the present study are available upon reasonable request to the authors

## Data availability

Data is available upon a reasonable request to the corresponding author, and upon approval of the corresponding Data Transfer Agreement.

## Acknowledgements

**EGG, AC, MA, EMH, LB** have nothing to disclose. **NMO** had a predoctoral Rio Hortega fellowship, from the Instituto de Salud Carlos III (CM21/00018); has been awarded an ECTRIMS-MAGNIMS Research Fellowship 2024; also received speaking honoraria and travel expenses from Merck and Roche. **AVJ** has received research support from Fondo de Investigación en Salud (PI17/02162 and PI22/01589) from Instituto de Salud Carlos III, Spain, and in the last 12 months has engaged in consulting and/or participated as a speaker in events organised by Roche, Novartis, Merck, Neuraxpharm, and Lundbeck, and is Editor-in-Chief of Revista Neurologia. **XM** Professor Montalban’s institution has received compensation for lecture honoraria and travel expenses, participation in scientific meetings, clinical trial steering committee membership, or clinical advisory board participation in recent years from Abbvie, Actelion, Alexion, Bial PD, Biogen, Bristol-Myers Squibb/Celgene, EMD Serono, Genzyme, Hoffmann-La Roche, Immunic Therapeutics, Janssen Pharmaceuticals, Medday, Merck, Mylan, Nervgen, Neuraxpharm, Novartis, Peervoice, Samsung-Biosys, Sandoz, Sanofi-Genzyme, Teva Pharmaceutical, TG Therapeutics, Excemed, Medscape, ECTRIMS, MSIF, and NMSS or any of their affiliates. **MT** received consulting compensation and speaker honoraria from Almirall, Bayer Schering Pharma, Biogen-Idec, Genzyme, Merck-Serono, Novartis, Roche, Sanofi-Aventis, Viela-Bio and Teva Pharmaceuticals. She also declared her role as co-editor of Multiple Sclerosis Journal—Experimental, Translational and Clinical. **AR** serves/ed on scientific advisory boards for Novartis, Sanofi-Genzyme, Synthetic MR, TensorMedical, Roche, and Biogen, and has received speaker honoraria from Bayer, Sanofi-Genzyme, Merck-Serono, Teva Pharmaceutical Industries Ltd, Novartis, Roche, Bristol-Myers and Biogen, is CMO and co-founder of TensorMedical, and receives research support from Fondo de Investigación en Salud (PI19/00950 and PI22/01589) from Instituto de Salud Carlos III, Spain. **JS-G** receives research support from the Instituto de Salud Carlos III, co-funded by the European Union (PI19/0950, PI22/0750, PI25/01664), has participated in events funded by Merck, Roche, BMS, Sanofi, Novartis, Neuraxpharm, and Cobel-Darou in the last 36 months, and is a member of the ECTRIMS Executive Board and Editor-in-Chief of the Multiple Sclerosis Journal. **DP** receives research support from the Instituto Carlos III, cofunded by the European Union (PI18/00823, PI22/01709).

## Authors contributions

EGG and DP had full access to all the data in the study and take responsibility for the integrity of the data and the accuracy of the data analysis. Concept and design: EGG, ACL, JS-G, DP. Data acquisition and cohort curation: MA, LB, NMO, RMH, AVJ, XM, MT, AR, JS-G, DP. MRI processing and lesion analysis: EGG, ACL, MA. Radiomic analysis and machine-learning methodology: EGG, ACL, DP. Statistical analysis: EGG, ACL, DP. Interpretation of data: EGG, ACL, MA, LB, NMO, RMH, AVJ, XM, MT, AR, JS-G, DP. Drafting of the manuscript: EGG. Critical revision of the manuscript for important intellectual content: ACL, MA, LB, NMO, RMH, AVJ, XM, MT, AR, JS-G, DP. Supervision: JS-G, DP.

