## Supplementary_material for "Assessment of Dissemination in Time in Multiple Sclerosis Using a Single 3D T2-FLAIR magnetic resonance imaging Radiomic Signature"

Supplementary Figure 1. Cohort characteristics, longitudinal sampling, and lesion temporal categorisation

Supplementary Figure 1 provides the cohort-level descriptive context for the study. Age and sex distributions were broadly comparable across the three analysis groups, supporting the independence of the feature-selection, training-validation, and test cohorts without major demographic imbalance. The figure also shows the relative proportions of LS-E, R-I, and excluded lesions across groups, confirming that the temporal lesion composition was preserved despite the cohort split. Illustrative examples further clarify the temporal criteria used to assign lesions to LS-E and R-I categories and to exclude intermediate-duration lesions.

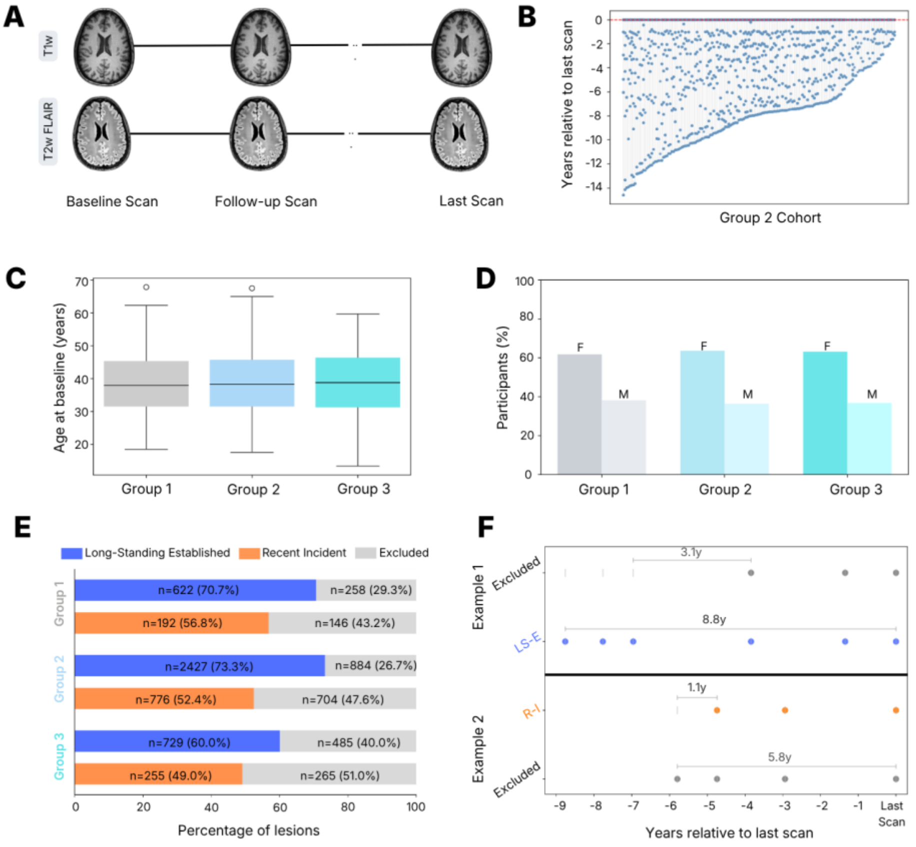


**Supplementary Figure 1. Cohort characteristics, longitudinal sampling, and lesion temporal categorisation. (A-B)** Distribution of longitudinal MRI sampling across cohorts, shown as years relative to the last available scan. **(C)** Age distribution at baseline across Group 1, Group 2, and Group 3. **(D)** Sex distribution across groups. **(E)** Proportion of lesions classified as LS-E, R-I, or excluded in each group. **(F)** Schematic representation of temporal lesion categorisation, illustrating assignment of lesions to LS-E (>6 years), R-I (<2 years), or excluded intermediate-duration categories.

Supplementary Figure 2. Cross-validation and final-model performance in the training cohort

Supplementary Figure 2 summarises classifier performance during model development in Group 2. Repeated cross-validation showed stable discrimination between lesion classes, with a mean AUC of approximately 0.81 and broadly consistent class-wise performance across folds. Performance of the final model trained on the full Group 2 cohort was similar, supporting the robustness of the selected radiomic feature set and the stability of the learned decision boundary before independent testing.

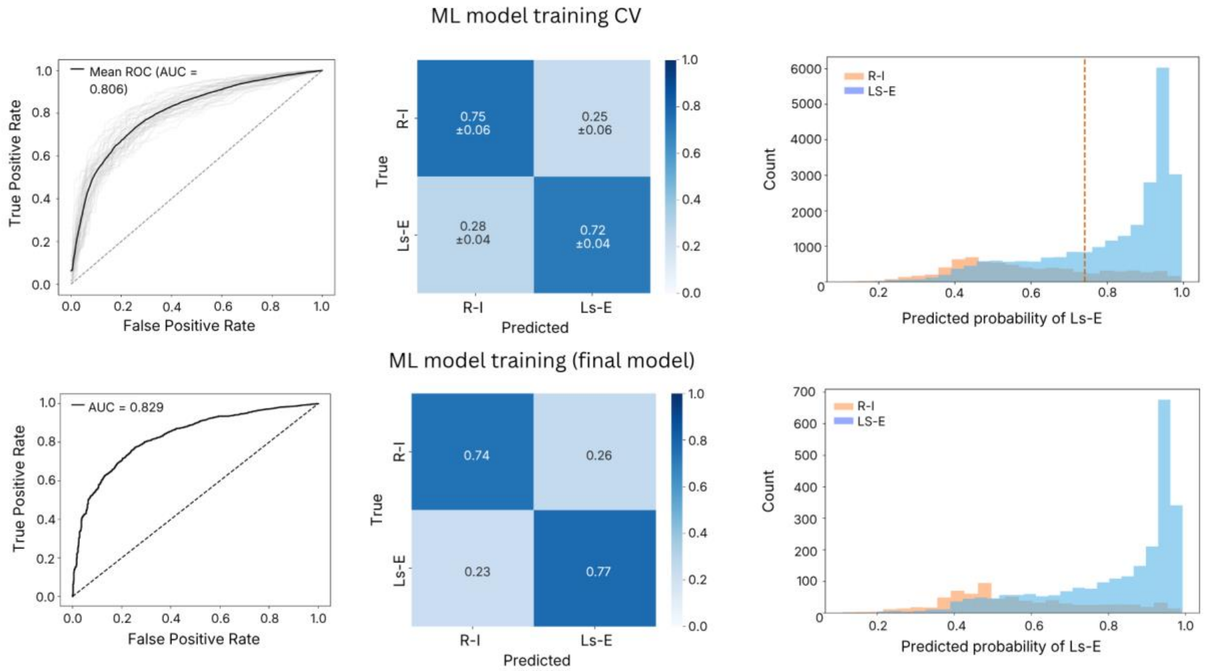


**Supplementary Figure 2. Cross-validation and final-model performance in the training cohort.** ROC curves, confusion matrices, and predicted-probability distributions for the Group 2 training cohort. Top row: repeated cross-validation performance, including mean ROC curve, average confusion matrix, and pooled predicted-probability distributions. Bottom row: performance of the final model trained on the full Group 2 dataset. These analyses show consistent discrimination between LS-E and R-I lesions during model development.

<u>Supplementary Figure 3. Temporal sampling windows in the discovery and test cohorts</u>

Supplementary Figure 3 illustrates the temporal distribution of MRI time points relative to the last available scan in Groups 1 and 3. This highlights the heterogeneous but clinically realistic retrospective follow-up structure of the dataset and shows that both the feature-discovery and independent test cohorts spanned a broad temporal sampling window. These distributions support the feasibility of defining temporally separated lesion categories in real-world longitudinal MRI data.

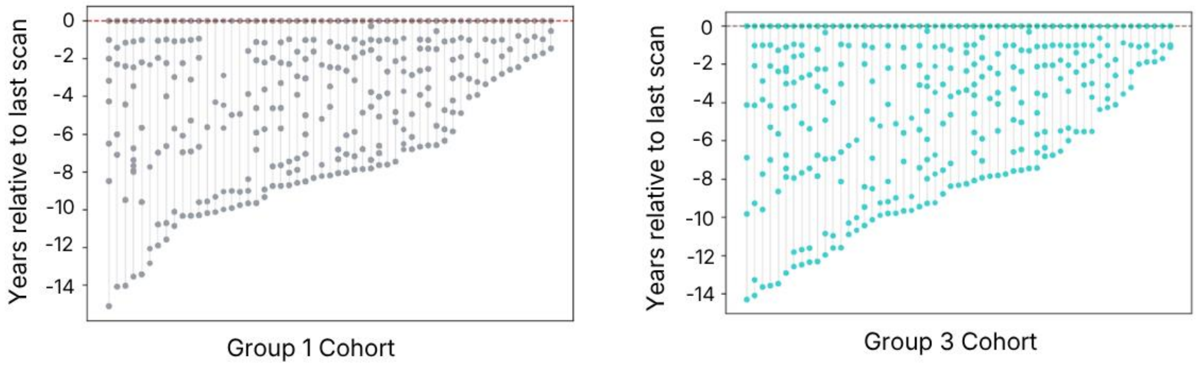


**Supplementary Figure 3. Temporal sampling windows in the discovery and test cohorts.** Distribution of MRI acquisitions relative to the last available scan in Group 3 and Group 1. Each point represents one MRI time point, shown in years before the last scan. These panels illustrate the heterogeneous longitudinal sampling available for temporal lesion stratification in the independent test and feature-selection cohorts.

**Supplementary Table 1. Sensitivity of LS-E vs R-I classification to radiomic feature redundancy**

| Strategy | Inputs | G2 repeated grouped-CV AUC | Delta G2 AUC | G2 participant-weighted AUC | G3 independent-test AUC | Delta G3 AUC |
|---|---|---|---|---|---|---|
| **All 18 selected features** | 18 | 0.806 +/- 0.030 | +0.000 | 0.809 | 0.778 | +0.000 |
| Correlation-pruned \|r\| > 0.90 | 9-10 | 0.802 +/- 0.032 | -0.004 | 0.803 | 0.769 | -0.009 |
| Correlation-pruned \|r\| > 0.85 | 8 | 0.803 +/- 0.031 | -0.003 | 0.805 | 0.768 | -0.009 |
| Correlation-pruned \|r\| > 0.80 | 5 | 0.773 +/- 0.030 | -0.033 | 0.770 | 0.743 | -0.035 |
| Intensity only | 6 | 0.793 +/- 0.030 | -0.013 | 0.796 | 0.762 | -0.016 |
| Geometry only | 9 | 0.715 +/- 0.038 | -0.091 | 0.728 | 0.684 | -0.094 |
| Run-length texture only | 3 | 0.684 +/- 0.038 | -0.122 | 0.703 | 0.663 | -0.115 |
| Intensity + geometry | 15 | 0.801 +/- 0.029 | -0.005 | 0.801 | 0.773 | -0.005 |
| Intensity + texture | 9 | 0.802 +/- 0.030 | -0.005 | 0.807 | 0.774 | -0.004 |
| Geometry + texture | 12 | 0.725 +/- 0.038 | -0.081 | 0.740 | 0.702 | -0.075 |
| PCA retaining 95% variance | 5 | 0.751 +/- 0.034 | -0.055 | 0.754 | 0.734 | -0.043 |

Notes: All models used an SVM (kernel='rbf', C=1, gamma='scale', class_weight='balanced'). G2 performance is the mean of 50 held-out participant-grouped folds (5 folds x 10 repetitions). Correlation pruning was performed within each G2 training fold; for G3, the retained feature set was derived from all G2 data before fitting and was then applied unchanged to G3. G3 was not used for feature selection. AUC differences are descriptive sensitivity analyses, not equivalence tests.

**Supplementary Table 2. Sensitivity analysis for unequal lesion contribution across participants**

| Model | Training weighting | G2 CV lesion AUC | G2 participant-weighted AUC | G3 lesion AUC (95% CI) | G3 participant-weighted AUC (95% CI) | LS-E sensitivity | R-I specificity |
|---|---|---|---|---|---|---|---|
| **Reference SVM** | Class-balanced, equal weight per lesion | **0.806** | **0.809** | **0.778 (0.739–0.813)** | **0.773 (0.722–0.822)** | 73.5% | 65.1% |
| **Participant-weighted SVM** | Inverse participant lesion count + class balancing | **0.797** | **0.798** | **0.767 (0.724–0.806)** | **0.760 (0.708–0.810)** | 70.7% | 71.4% |

Notes: Participant weighting assigned each lesion an initial weight inversely proportional to the number of lesions contributed by that participant, thereby reducing the influence of participants with high lesion burden. Weights were recalculated within each training subset and subsequently combined with class balancing. Confidence intervals were obtained by bootstrap resampling participants with all corresponding lesions retained.

**Supplementary Table 3. Paired radiomic feature differences in Group 1**

| Domain | Feature | Paired Cohen d | Direction | Unadjusted P | FDR-adjusted P |
|---|---|---|---|---|---|
| First-order | Maximum | 1.65 | Higher in LS-E | 1.56e-10 | 1.60e-08 |
| Shape | Sphericity | -1.30 | Higher in R-I | 2.95e-08 | 1.08e-06 |
| Shape | Major Axis Length | 1.29 | Higher in LS-E | 3.15e-08 | 1.08e-06 |
| Shape | Maximum 3D Diameter | 1.26 | Higher in LS-E | 4.97e-08 | 1.28e-06 |
| First-order | Mean | 1.18 | Higher in LS-E | 1.72e-07 | 3.26e-06 |
| Shape | Maximum 2D Diameter, Row | 1.18 | Higher in LS-E | 1.94e-07 | 3.26e-06 |
| Shape | Flatness | -1.17 | Higher in R-I | 2.21e-07 | 3.26e-06 |
| First-order | Root Mean Squared | 1.14 | Higher in LS-E | 3.38e-07 | 4.36e-06 |
| GLRLM | Run Variance | 1.11 | Higher in LS-E | 5.54e-07 | 6.34e-06 |
| Shape | Maximum 2D Diameter, Slice | 1.09 | Higher in LS-E | 7.61e-07 | 7.84e-06 |
| Shape | Maximum 2D Diameter, Column | 1.08 | Higher in LS-E | 8.65e-07 | 7.91e-06 |
| First-order | 90th Percentile | 1.08 | Higher in LS-E | 9.21e-07 | 7.91e-06 |
| First-order | 10th Percentile | 1.06 | Higher in LS-E | 1.19e-06 | 9.44e-06 |
| Shape | Minor Axis Length | 0.97 | Higher in LS-E | 5.59e-06 | 4.11e-05 |
| GLRLM | Run Length Non-Uniformity Normalized | -0.93 | Higher in R-I | 9.93e-06 | 6.82e-05 |
| Shape | Elongation | -0.91 | Higher in R-I | 1.30e-05 | 8.40e-05 |
| First-order | Median | 0.89 | Higher in LS-E | 1.88e-05 | 1.14e-04 |
| GLRLM | Run Entropy | 0.83 | Higher in LS-E | 5.11e-05 | 2.92e-04 |

**Notes:** Feature selection used 32 Group 1 participants who contributed both lesion categories. Lesion-level features were averaged within participant and lesion category before paired testing, yielding 32 paired observations per feature. Cohen d is the paired effect size; positive values indicate higher values in LS-E lesions and negative values indicate higher values in R-I lesions. False discovery rate adjustment used the Benjamini-Hochberg procedure. All 18 features shown met both prespecified criteria: FDR-adjusted P < .05 and absolute Cohen d > 0.8. GLRLM indicates gray-level run-length matrix.

**Supplementary Table 4. Held-out test performance with participant-cluster bootstrap 95% CIs**

| Metric | Numerator/denominator | Estimate | 95% CI |
|---|---|---|---|
| Area under the ROC curve | NA | 0.778 | 0.739-0.814 |
| LS-E sensitivity | 457/622 | 73.5% | 68.5%-77.9% |
| R-I specificity | 125/192 | 65.1% | 57.0%-73.5% |
| Accuracy | 582/814 | 71.5% | 67.5%-75.2% |
| Balanced accuracy | NA | 0.693 | 0.647-0.737 |
| LS-E positive predictive value | 457/524 | 87.2% | 80.9%-91.4% |
| R-I negative predictive value | 125/290 | 43.1% | 32.0%-56.5% |
| LS-E F1 score | NA | 0.798 | 0.754-0.829 |
| R-I F1 score | NA | 0.519 | 0.420-0.622 |
| Brier score | NA | 0.152 | 0.129-0.188 |

**Notes:** The held-out test cohort contained 814 lesions from 55 participants (622 LS-E and 192 R-I). Confidence intervals were estimated with 5,000 participant-cluster bootstrap resamples, sampling participants with replacement and retaining all lesions contributed by each sampled participant. The fixed decision threshold was selected in Group 2 and applied unchanged to Group 3. LS-E was treated as the positive class. ROC indicates receiver operating characteristic.

**Sensitivity-analysis result to retain separately:** participant-weighted AUC, 0.773 (95% CI, 0.720-0.822).

STROBE Statement—Checklist of items that should be included in reports of ***cohort studies***

| | Item No | Recommendation | Page No |
|---|---|---|---|
| **Title and abstract** | 1 | (*a*) Indicate the study's design with a commonly used term in the title or the abstract | 1 |
| | | (*b*) Provide in the abstract an informative and balanced summary of what was done and what was found | 1 |
| **Introduction** | | | |
| Background/rationale | 2 | Explain the scientific background and rationale for the investigation being reported | 2 |
| Objectives | 3 | State specific objectives, including any prespecified hypotheses | 2-3 |
| **Methods** | | | |
| Study design | 4 | Present key elements of study design early in the paper | 3 |
| Setting | 5 | Describe the setting, locations, and relevant dates, including periods of recruitment, exposure, follow-up, and data collection | 3 |
| Participants | 6 | (*a*) Give the eligibility criteria, and the sources and methods of selection of participants. Describe methods of follow-up | 3-4 |
| | | (*b*) For matched studies, give matching criteria and number of exposed and unexposed | N/A |
| Variables | 7 | Clearly define all outcomes, exposures, predictors, potential confounders, and effect modifiers. Give diagnostic criteria, if applicable | 4-6 |
| Data sources/ measurement | 8* | For each variable of interest, give sources of data and details of methods of assessment (measurement). Describe comparability of assessment methods if there is more than one group | 3-6 |
| Bias | 9 | Describe any efforts to address potential sources of bias | 4-5 |
| Study size | 10 | Explain how the study size was arrived at | 3 |
| Quantitative variables | 11 | Explain how quantitative variables were handled in the analyses. If applicable, describe which groupings were chosen and why | 3-6 |
| Statistical methods | 12 | (*a*) Describe all statistical methods, including those used to control for confounding | 4-6; Suppl 4-6 |
| | | (*b*) Describe any methods used to examine subgroups and interactions | 5-7 |
| | | (*c*) Explain how missing data were addressed | 15 |
| | | (*d*) If applicable, explain how loss to follow-up was addressed | N/A; 3 |
| | | (*e*) Describe any sensitivity analyses | 5-7; Suppl 4-6 |
| **Results** | | | |
| Participants | 13* | (a) Report numbers of individuals at each stage of study—eg numbers potentially eligible, examined for eligibility, confirmed eligible, included in the study, completing follow-up, and analysed | 3, 6, 14-15 |
| | | (b) Give reasons for non-participation at each stage | 3-4 |
| | | (c) Consider use of a flow diagram | 12 |
| Descriptive data | 14* | (a) Give characteristics of study participants (eg demographic, clinical, social) and information on exposures and potential confounders | 3, 14-15 |
| | | (b) Indicate number of participants with missing data for each variable of interest | 15 |
| | | (c) Summarise follow-up time (eg, average and total amount) | 3, 14-15 |

| | | | |
|---|---|---|---|
| Outcome data | 15* | Report numbers of outcome events or summary measures over time | 6-7; Suppl 6 |
| Main results | 16 | (*a*) Give unadjusted estimates and, if applicable, confounder-adjusted estimates and their precision (eg, 95% confidence interval). Make clear which confounders were adjusted for and why they were included | 6; Suppl 6 |
| | | (*b*) Report category boundaries when continuous variables were categorized | 4-5 |
| | | (*c*) If relevant, consider translating estimates of relative risk into absolute risk for a meaningful time period | N/A |
| Other analyses | 17 | Report other analyses done—eg analyses of subgroups and interactions, and sensitivity analyses | 5-7; Suppl 4-6 |
| **Discussion** | | | |
| Key results | 18 | Summarise key results with reference to study objectives | 7, 10 |
| Limitations | 19 | Discuss limitations of the study, taking into account sources of potential bias or imprecision. Discuss both direction and magnitude of any potential bias | 9-10 |
| Interpretation | 20 | Give a cautious overall interpretation of results considering objectives, limitations, multiplicity of analyses, results from similar studies, and other relevant evidence | 7-10 |
| Generalisability | 21 | Discuss the generalisability (external validity) of the study results | 10 |
| **Other information** | | | |
| Funding | 22 | Give the source of funding and the role of the funders for the present study and, if applicable, for the original study on which the present article is based | 10 |

*Give information separately for exposed and unexposed groups.
